# Probing the dissonance among the diagnostic outputs of multiple approaches used for detection of Methicillin-resistant *Staphylococcus aureus* (MRSA)

**DOI:** 10.1101/2020.07.20.20158519

**Authors:** Ujjwal Ranjan Dahiya, Arnab Sikidar, Priyanka Sharma, Chitra Rawat, Benu Dhawan, Arti Kapil, Ravikrishnan Elangovan, Dinesh Kalyanasundaram

## Abstract

Methicillin resistant staphylococcus aureus (MRSA) is an extremely infectious hospital acquired bacterial pathogen often found in post-surgical patients globally. Early detection of such pathogens is a critical requirement to eliminate or reduce the incidence of anti-microbial resistance as well as for effective management of the disease. Despite the development of multiple biochemical, microbiological and nucleic acid amplifications techniques (NAATs), conventional culture methods are widely used clinically owing to high variability between the methods, technical skills and infrastructural needs. Further, multiple reports suggest significant variation among diagnostic output for MRSA detection. This work attempts to probe the discordance among the diagnostic output of three commonly used methods, while trying to understand the underlying cause of variability. MRSA detection on 217 clinical pus isolates was carried out using three different methods namely, conventional culture method, qPCR-based amplification and a modern LAMP based detection approach. Also, to confirm the presence of MRSA and distinguish from coagulase-negative staphylococci (CoNS), as well as to investigate the observed differences between qPCR and LAMP outputs, melt curve analysis was performed on discordant samples. LAMP based MRSA detection was found to be the optimum method. In summary, this study evaluates the diagnostic efficiency of the different detection methods, while probing for possible explanations for the observed differences.

## 1. Introduction

Globally, methicillin-resistant *Staphylococcus aureus* (MRSA) is associated with nosocomial and community-acquired healthcare infections (1,2). Due to the extensive use of antibiotics, an increasing number of antibiotic-resistant (ABR) bacterial strains such as MRSA is causing enormous healthcare challenge to mankind (3,4). The pathogen is capable of producing a diverse range of toxins and virulence factors, including toxic shock syndrome toxin (TSST) (5). A higher mortality rate of about twenty-two percent is observed in MRSA infected patients in comparison to five percent amongst non-MRSA patients (6–8). The resistance of MRSA towards β-lactam based antibiotics including methicillin is caused by the presence of inherent β-lactamase as well as the expression of *mecA* gene resulting in the production of penicillin-binding proteins (PBP, PBP2, and PBP2a) that shows lower affinity to β-lactam based antibiotics (9). Rapid identification and timely isolation of MRSA infected subjects are crucial to avoid complications (10,11). Conventional methods for the detection of MRSA can take up to 48 hours or more time due to time-consuming protocols including culture, colony morphology, and anti-microbial susceptibility testing (12,13). Lately, health institutions are giving high importance to rapid identification of bacterial isolates and screening of their antimicrobial susceptibility, especially in positive blood culture isolates (14,15). This led to deployment of multiple nucleic acid amplification techniques (NAAT) based approaches for fast detection of MRSA and other pathogens (16). Many commercial assays based on polymerase chain reaction (PCR) such as FluoroType® MRSA system (Hain-life science GmbH, Nehren, Germany) have been developed. Although these assays are costly and requires high technical skills (17,18).

Recently, the next step taken towards making MRSA detection fast and affordable with high sensitivity and selectivity is the use of isothermal amplification approaches (19,20). The various isothermal amplification methods are loop-mediated isothermal amplification (LAMP), primer-generation rolling circle amplification (PG-RCA), recombinase polymerase amplification (RPA), nucleic acid sequence-based amplification (NASBA), helicase-dependent amplification (HDA), exponential amplification reaction (EXPAR), and whole genome amplification (WGA). Among these, LAMP is a popular, well-studied, and standardized nucleic acid amplification technique with high specificity and sensitivity (21). LAMP utilizes polymerases such as Bst, capable of auto cycling strand-displacement mediated amplification. A set of 4 or 6 specific primers along with dNTPs are necessary for target sequence amplification (19). Amplification through LAMP method can be detected through multiple methods viz. turbidimetric, fluorescence and pH changes in the reaction mixture (22,23). Recent work from our group reported a portable system *SMOL* for the rapid diagnosis of *Salmonella* Typhi and *Salmonella* Paratyphi A (14).

While working towards designing a LAMP-based assay for MRSA detection, a significant discordance in diagnostic results was often observed in comparison to results of detection through culture method and qPCR. Perplexed with the observation, we looked in the literature and found multiple reports suggesting the differences in diagnostic results in MRSA detection (24–27). In the present work, we aim to understand the degree of and underlying factors behind such discordance. To achieve this, we compared the diagnostic results in 217 mixed flora clinical samples, obtained by three different approaches viz. LAMP-assay, qPCR method and culture method. The performance of these diagnostic methods is comprehensively compared with each other in terms of sensitivity and robustness. Available literature suggests presence of multiple pathogens, inhibitory protein and coagulase-negative staphylococci (CoNS) contamination as some of the putative source of error in MRSA detection (28). To assess the role of CoNS contamination, discordant results between NAAT methods were subjected to melt curve analysis (MCA). Mass spectrophotometry based detection of random samples was tried to substantiate the finding. The study designed is comprehensively illustrated in Figure 1. The manuscript while comprehensively assessing the effectiveness and robustness of these diagnostic methods, also highlight and discuss the critical factors to be considered while developing a rapid MRSA detection assay.

**Figure 1.**
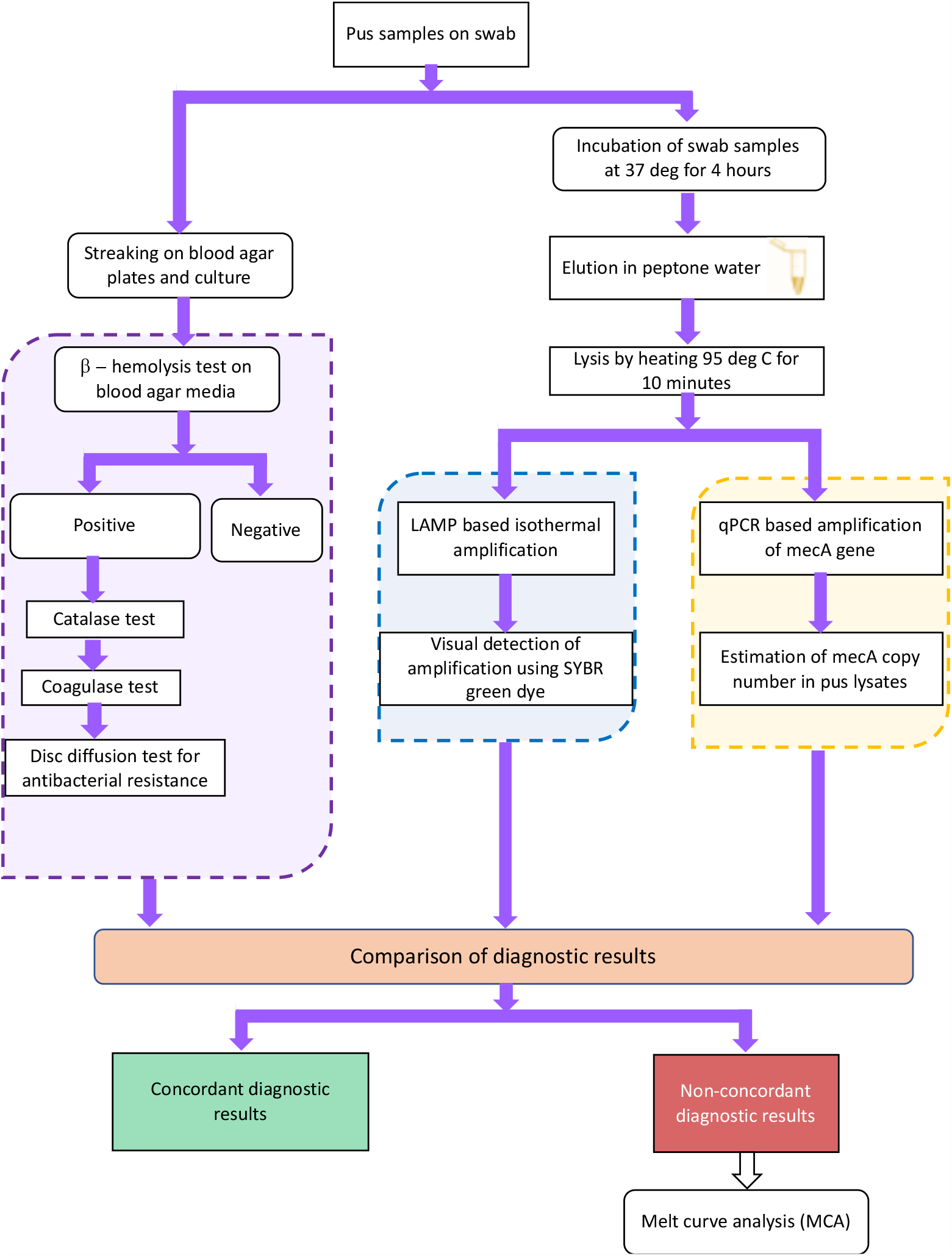
Schematic showing workflow for the comparative study.

## 2. Materials and methods

### 2.1 Institute ethical approval and collection of clinical samples

The study was ethically approved by Institutional ethics committee (document number IEC-569/02 dated 02.11.2018). Two hundred and seventeen clinical human pus samples were collected from 203 patients with clinical symptoms of *Staphylococcus aureus* infection. In fourteen patients, two samples were collected on different dates. The samples were collected after taking consents from the volunteering patients. The pus samples were sent to the department of microbiology from wards and out-patient department of All India Institute of Medical Sciences, New Delhi.

### 2.2 Culture-based detection of MRSA

The detailed protocol for the conventional culture-based diagnostic approach for MRSA is given in the supplementary section and involved culturing of the clinical samples on blood and MacConkey Agar plates followed by biochemical tests. Clinical pus aspirates were plated on blood Agar plate in carbon dioxide enriched atmosphere (5% CO_2_ incubator) for performing a β-hemolysis test. The presence of yellow to cream white colonies signified the presence of *S. aureus* or *Streptrococcus pyrogens*. For differentiating between two possible pathogens, catalase tests were performed by mixing a small volume of inoculum from each sample to a 3 % hydrogen peroxide solution. The release of bubbles confirms the presence of MRSA in clinical isolates. Further, coagulase tests were performed by incubating the pus samples with coagulase plasma (HIMEDIA®, India) for observation of blood clots on a glass slide. Cefoxitin was used as a marker to detect methicillin resistance through antibiotic disc diffusion tests and the results were analyzed by current CLSI guidelines.(29) To understand the limit of detection, serial dilutions of MRSA cells spiked blood culture media were used for standardizations for concentration range of 5 to 500 CFU/mL (supplementary section). The conventional culture method takes about 72 hours. All the culture-based detection of clinical pus samples were carried out at the Department of Microbiology of the associated hospital.

### 2.3 Sample processing for NAAT based detection

Elution of samples in buffer was performed post 4 hours of incubation of the pus samples, followed by lysis at 95 °C for 10 minutes in heated water bath. Pus sample supernatant thus prepared was stored at 4 °C until required further. Supernatant from each clinical sample was then used for setting up LAMP or qPCR reactions.

### 2.4 LAMP based detection of MRSA

LAMP reagents master mix were purchased from Optigene Private Limited, India, a subsidiary of Ampligene, UK. The primers for *mecA* gene were custom designed by our group and synthesized by Integrated DNA Technologies, USA, for LAMP experiments. LAMP based nucleic acid amplification used in this study was based on 4 primers; forward outer primer (F3), backward outer primer (B3), forward inner primer (FIP), and backward inner primer (BIP). The FIP, BIP, F3, and B3 primers for the *mecA* gene were designed in PrimerExplorer® version 5 (Table 1). BLAST® program was used for verification of primer specificity, prior to experimentation. The LAMP method has been explained elsewhere in detail.(30) Before setting up the reactions, solutions for primer mix and LAMP reagents mix were prepared to enable faster, convenient and precise experimentation. For preparing primer mix, 20 µL from 100 µM of FIP and BIP each, 5 µL from 100 µM of F3 and B3 each, and 30 µL of nuclease free water were mixed. For the LAMP assay, 5 µL of primer mix, 10 µL of LAMP reagent mix, 5 µL of clinical sample lysate (template DNA) and 5 µL of nuclease free water was used to setup a 25 µL reaction. All the samples were placed in a vial and sealed with parafilm before the treatment at 65 °C for LAMP. Amplified LAMP reactions were tested after adding 2 µL of 1000X SYBR green in each tube, followed by visual detection of positive samples.

**Table 1.**
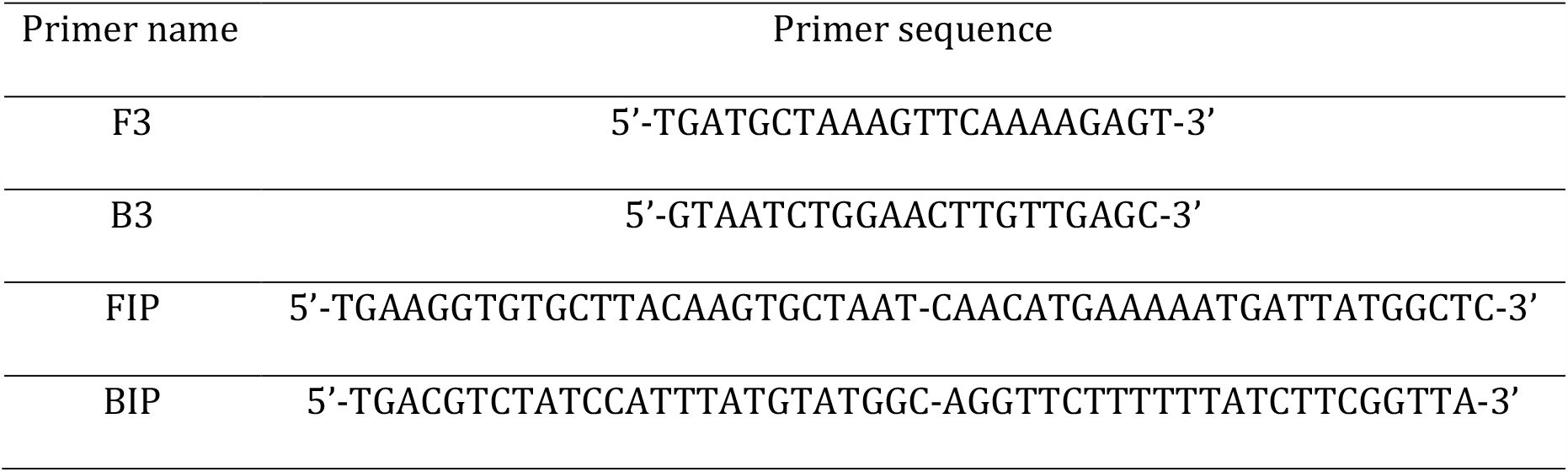
Sequence of primers used for LAMP based amplification of *mecA* gene.

### 2.5 Cross-reactivity and specificity of LAMP assay

Several common pathogenic bacterial species such as *S*. Havana, *S*. Paratyphi B, *S*. Typhimurium, *S*. Typhi, *Escherichia coli, Pseudomonas aeruginosa, Acinetobacter baumanii, Enterococcus fecalis, Klebsiella pneumonia, and Shigella flexneri* at a concentration of 10^6^CFU/mL were tested individually to evaluate the cross-reactivity of the primers. Similar protocol of DNA isolation and LAMP amplifications were followed as described in the preceding sections.

### 2.6 Quantitative *polymerase chain reaction* (*q*PCR)

KAPA SYBR master mix 2X was procured from Sigma Aldrich, USA, for *q*PCR-based amplifications. All other reagents including buffers, broth, albumin, etc., were obtained from Sigma-Aldrich, USA. qPCR reactions were carried out for validation of the results of clinical samples. In addition to the above, known dilution of MRSA culture was used to generate the standard curve between the *mecA* copy numbers (considering one copy of the gene per CFU) and *C*_t_ values (obtained in the *q*PCR reactions). The outer primers F3 and B3 were used in qPCR reactions to amplify the *mecA* gene. The standard curve of *C*_t_ vs. copy number was used for estimating the copy number in the clinical pus samples. The lysate prepared (described in the earlier sections) was used for the *q*PCR reactions. For both the clinical samples and for the standard curve experiments, 7.5 µL of KAPA SYBR 2X (Promega, USA) master mix, 2 µL of clinical sample lysate and 3 µL of pre-mixed primers (10 μM of F3 and B3 each) were used. The reactions were carried out at 95°C for 30 s for initial denaturation followed by 40 cycles of denaturation at 95°C for 10 s, and extension at 57°C for 30 s. The reactions were carried out in real time qPCR machine LightCycler® 480 (Roche molecular systems Inc., USA).

### 2.7 Melt Curve Analysis for differentiation MRSA from CoNS

To further probe the differences observed between the diagnostic methods, melt curve analysis (MCA) was performed. Primer sequences (Table 2) specific to the conservative domains of *Staphylococcus* genus (*16S*), *S. aureus* (*ITS*), and *mecA* gene were used for the assay.(31) For MCA assay, qPCR reactions were setup to check amplification of *16S, ITS*, and *mecA* genes in the samples with discordant results between the two NAAT diagnostic methods. The reactions were carried out in real-time *q*PCR machine CFX96® (BioRad Technologies, USA). Samples in which all three gene sequences were amplified were designated as MRSA, while samples with no amplification of *ITS* amplicon were designated as CoNS positive.

**Table 2.**
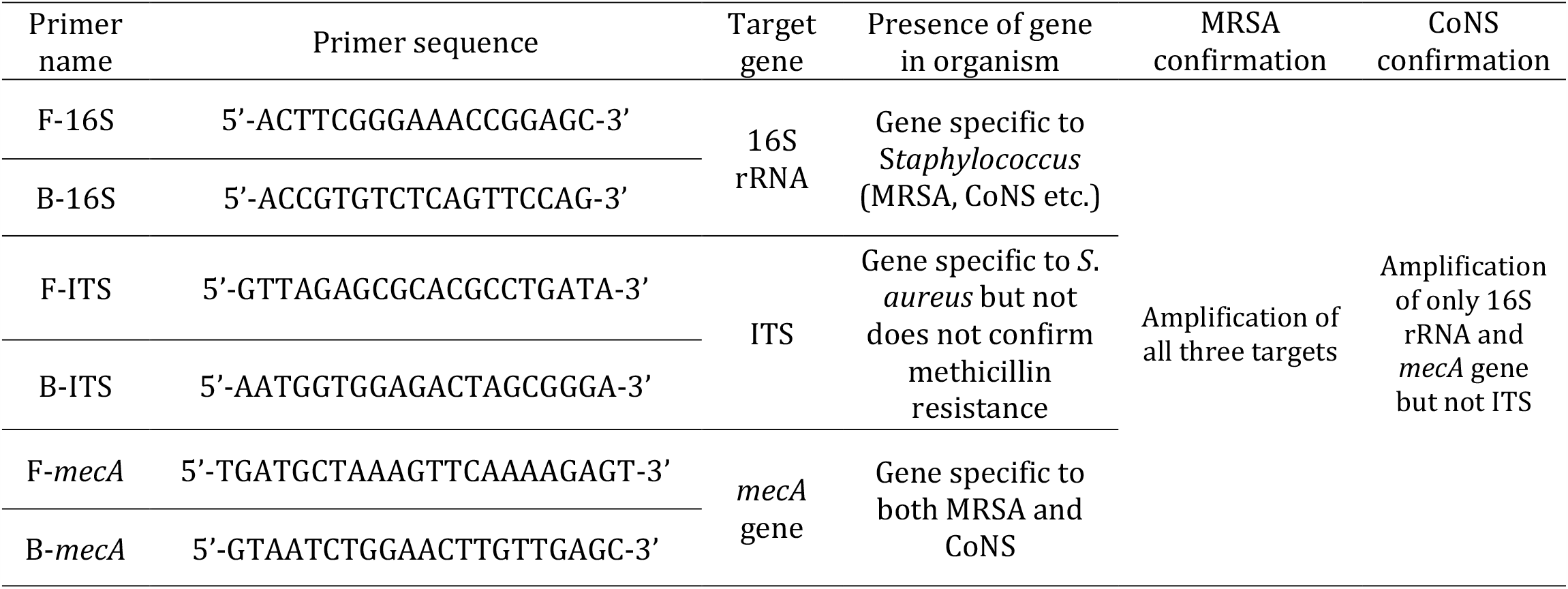
Primer sequence used for Melt Curve Analysis (MCA) based distinction between MRSA and CoNS.

### 2.8 Statistical analysis

The effectiveness and efficiency of the three diagnostic methods was estimated based on the diagnostic outputs, by calculating the clinical sensitivity and negative predictive value (NPV).(32) Power analysis was also performed on the results. For the sake of statistical analysis, the samples with negative results in all three methods (absence of MRSA) were taken as reference for estimating NPV and sensitivity.

## 3.0 Results

### 3.1 Cross-reactivity tests for the primers against other pathogens

The cross-reactivity was tested for the *mecA* primer sequences used for MRSA detection in the study. LAMP was performed against nucleic acids of the following bacterial pathogens: *S*. Havana, *S*. Paratyphi B, *S*. Typhimurium, *S*. Typhi, *Escherichia coli, Pseudomonas aeruginosa, Acinetobacter baumanii, Enterococcus fecalis, Klebsiella pneumonia, and Shigella flexneri*. Amplification in all samples was observed visually post addition of SYBR green. The primer sequences were found to be specific and no amplification was observed with DNA of above-mentioned organisms (Figure 2).

**Figure 2.**
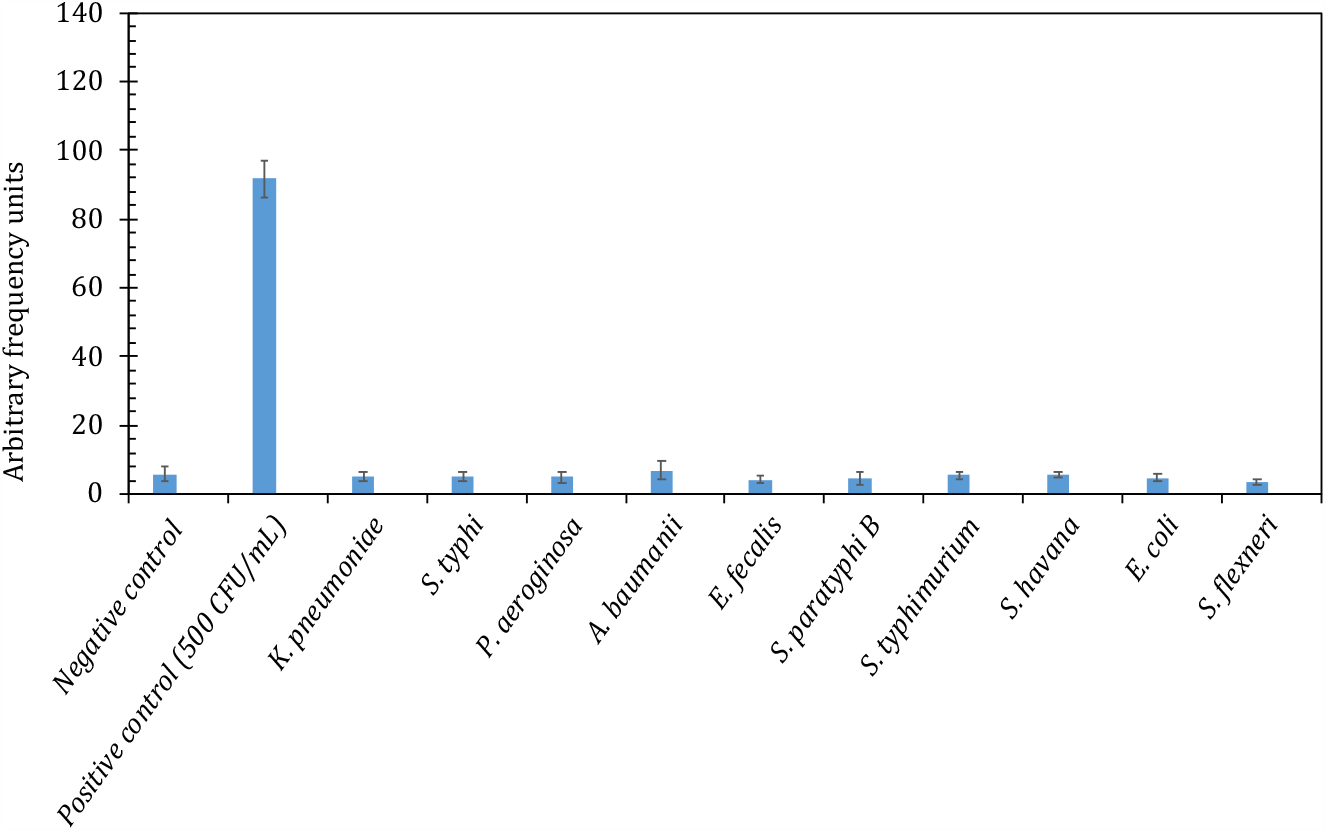
(a) Case wise comparison of diagnostic efficacy different molecular diagnostic methods used for MRSA detection. (b) Comparative performance of different diagnostic methods on 54 Clinical pus samples (6 samples showed negative results in all three methods while 48 clinical pus isolates recorded non-concordant results).

### 3.2 Diagnostic results for different approaches used for MRSA detection

MRSA detection on all 217 clinical samples was performed using threes diagnostic approaches as mentioned above and the diagnostic outputs were tabulated (Table 3). Conventional culture-based MRSA detection on 217 clinical samples resulted in 30 positive samples and 187 negatives (Figure 3). The isothermal amplification based LAMP method was confirmed using visual detection after adding SYBR green dye and showed 104 positive samples and 113 negatives.

**Table 3.**
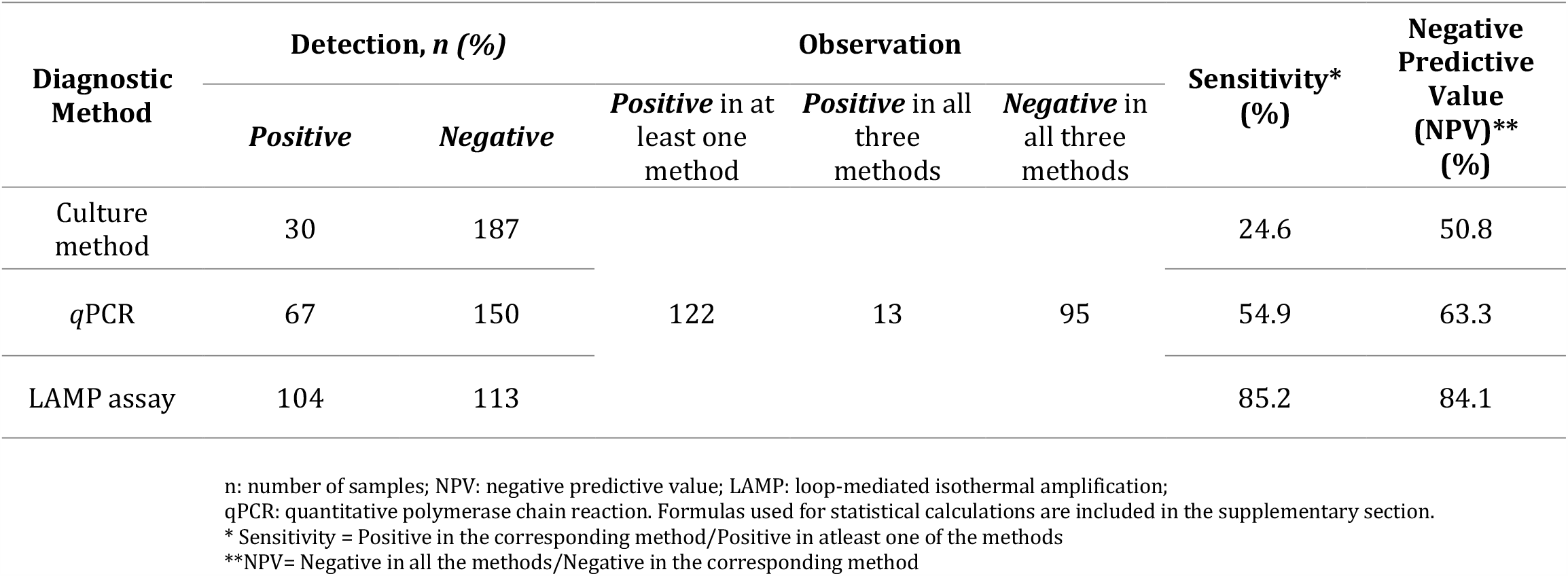
Culture results of the 225 clinical pus samples tested for MRSA presence using three different diagnostic methods. Statistical Analysis of diagnostic performance of the methods is included in the table.

**Figure 3.**
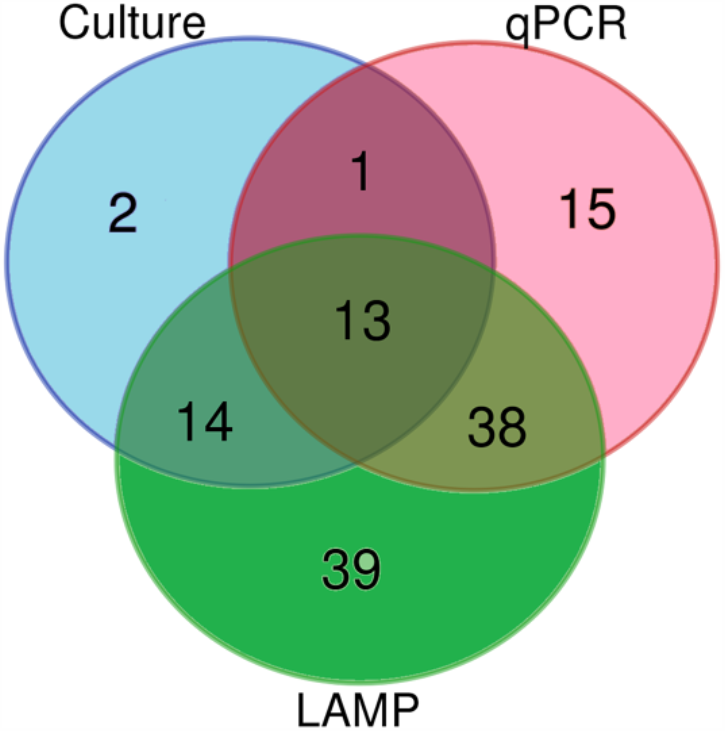
Comparative performance of different diagnostic methods on 122 positive clinical pus samples out of 217 total samples (95 samples showed negative results by all three methods and hence excluded from the diagram).

*q*PCR based diagnosis as well as quantification of *mecA* copy number in clinical pus samples was performed after preparing a standard curve between *C*_t_ vs *mecA* copy numbers (supplementary section). qPCR based estimation of *mecA* copy number showed distinct variation among the tested samples (supplementary section). Based on the standard curve, a cut off value of C_t_ equal to 23.65 was used for distinguishing between the positive and negative MRSA detection and further statistical analysis. qPCR based MRSA detection resulted in 67 positive samples and 150 negative samples (Figure 2). On comparing the diagnostic outputs, a total of 13 samples were found positive in all three methods while 95 samples were found negative across the methods (Figure 3). Sample-wise detailed results for all three diagnostic approaches are included in the supplementary tables.

### 3.3 Statistical significance of different approaches

The sensitivity and Net predictive values (NPV) for MRSA detection in three methods were estimated using statistical analysis based on the diagnostic outputs (Table 3). A total of 95 samples showed MRSA absence in all three approaches, and thus used as reference for estimating sensitivity and negative predictive value (NPV). Culture based MRSA detection showed a low sensitivity of 24.6% and NPV of 50.8%. LAMP based detection resulted in 85.2% sensitivity and 84.1% NPV. Sensitivity and NPV of 54.9% and 63.3% respectively was observed in the case of qPCR-based MRSA estimation.

### 3.4 Estimating the discordance in diagnostic outputs

Observed MRSA detection result using three different methods showed significant differences. To assess the degree of variability between the diagnostic outputs, we estimated percentage discordance (non-concordant as a percentage of total) within the results. The diagnostic outputs were categorized in 8 different group based on the results (Table 4a). While considering results from all three methods together, a total of 108 samples (13 positive and 95 negative) showed concordant results, resulting in overall discordance of 50.2%. Next, we estimated the pairwise discordance between the approaches (Table 4b). In pairwise comparison, highest discordance of 36.9% was observed between the results of culture and LAMP methods. This was followed by an estimated discordance of 31.8% between culture and qPCR results. Finally, between the two NAAT methods the estimated discordance was found to be 31.8%.

**Table 4a.** Estimating the discordance between the MRSA detection results through three different approaches. Diagnostic output of 217 clinical samples was categorized in 8 groups.

| Concordance | LAMP assay | qPCR | Culture method | n (%) |
| --- | --- | --- | --- | --- |
| \$Concordant | + | + | + | 13 |
| Discordant | + | + | - | 38 |
| Discordant | + | - | + | 14 |
| Discordant | - | + | + | 1 |
| Discordant | - | - | + | 2 |
| Discordant | - | + | - | 15 |
| Discordant | + | - | - | 39 |
| *Concordant | - | - | - | 95 |
| **Total** |  |  |  | 217 |
| **% Discordance = [(Total – Concordant)/Total]\*100** |  |  |  | 50.2 |

n: number of samples; NPV: negative predictive value; LAMP: loop-mediated isothermal amplification;
qPCR: quantitative polymerase chain reaction
\*True negative => Negative in all methods = 95
\$True positive => Positive in all methods = 13

**Table 4b.** Pairwise comparison and estimation of discordance in MRSA detection results using three different methods.

| **Concordance** | **Findings** | **Culture vs.**<br>**qPCR**<br>**n (%)** | **Culture vs.**<br>**LAMP**<br>**n (%)** | **qPCR vs.**<br>**LAMP**<br>**n (%)** |
| --- | --- | --- | --- | --- |
| Concordant | (+), (+) | 14 | 27 | 51 |
| Concordant | (-), (-) | 134 | 110 | 97 |
| Discordant | (+), (-) | 16 | 3 | 16 |
| Discordant | (-), (+) | 53 | 77 | 53 |
| **Total** |  | 217 | 217 | 217 |
| **% Discordance** = [(Total – Concordant)/Total]*100 |  | 31.8 | 36.9 | 31.8 |

n: number of samples; LAMP: loop-mediated isothermal amplification; qPCR: quantitative polymerase chain reaction

### 3.5 Role of CoNS presence in the mixed culture (Melt curve Analysis)

Since both qPCR and LAMP based detection of MRSA in the study was based on mecA gene, a significant discordance of 31.8% was poorly understood. To assess the role of CoNS contamination in the observed discordance we performed MCA. The 69 discordant samples between LAMP and qPCR methods were subjected to melt curve analysis (MCA) to find the presence of CoNS in the samples. Samples were tested for specific amplification peaks of mecA (79.5°C), ITS (86.5°C) and 16s rRNA (83.5°C) in melt curves. Amplification of ITS gene along with the mecA gene and 16S rRNA gene confirms the presence of MRSA, while absence of ITS peak in melt curve signified CoNS contamination. Melt curve analysis confirmed the presence of CoNS in 17 out of the 69 tested samples, while rest were found to be CoNS negative (Figure 4, Table 5).

**Table 5.**
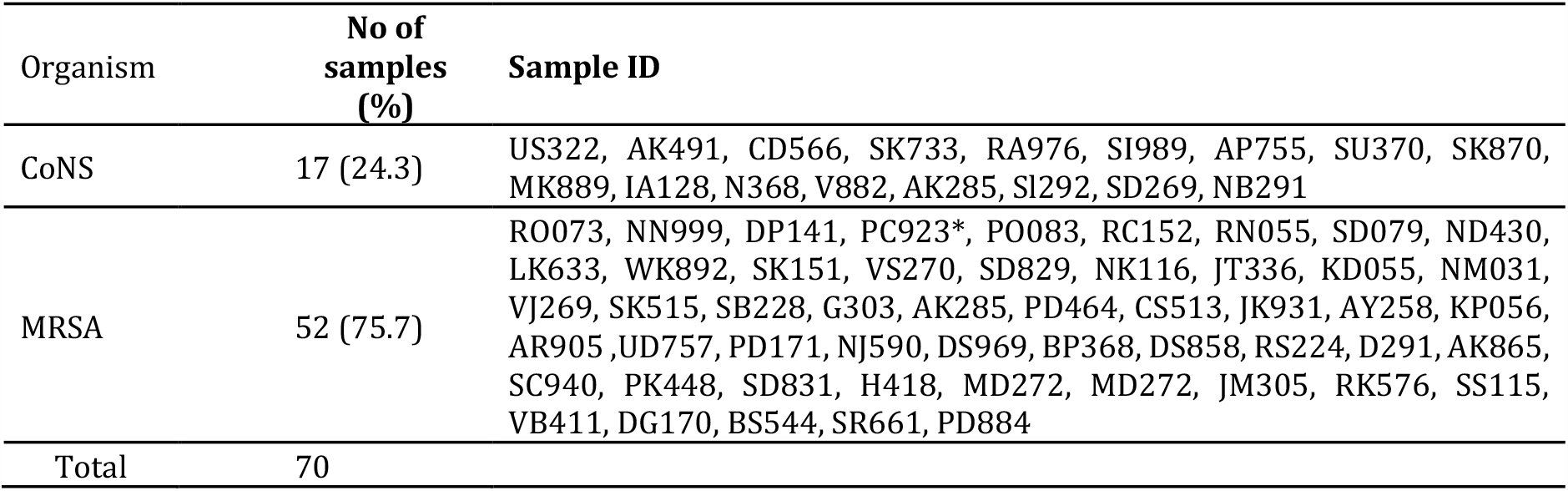
Summarized results for melt curve analysis for 69 discordant samples.

**Figure 4.**
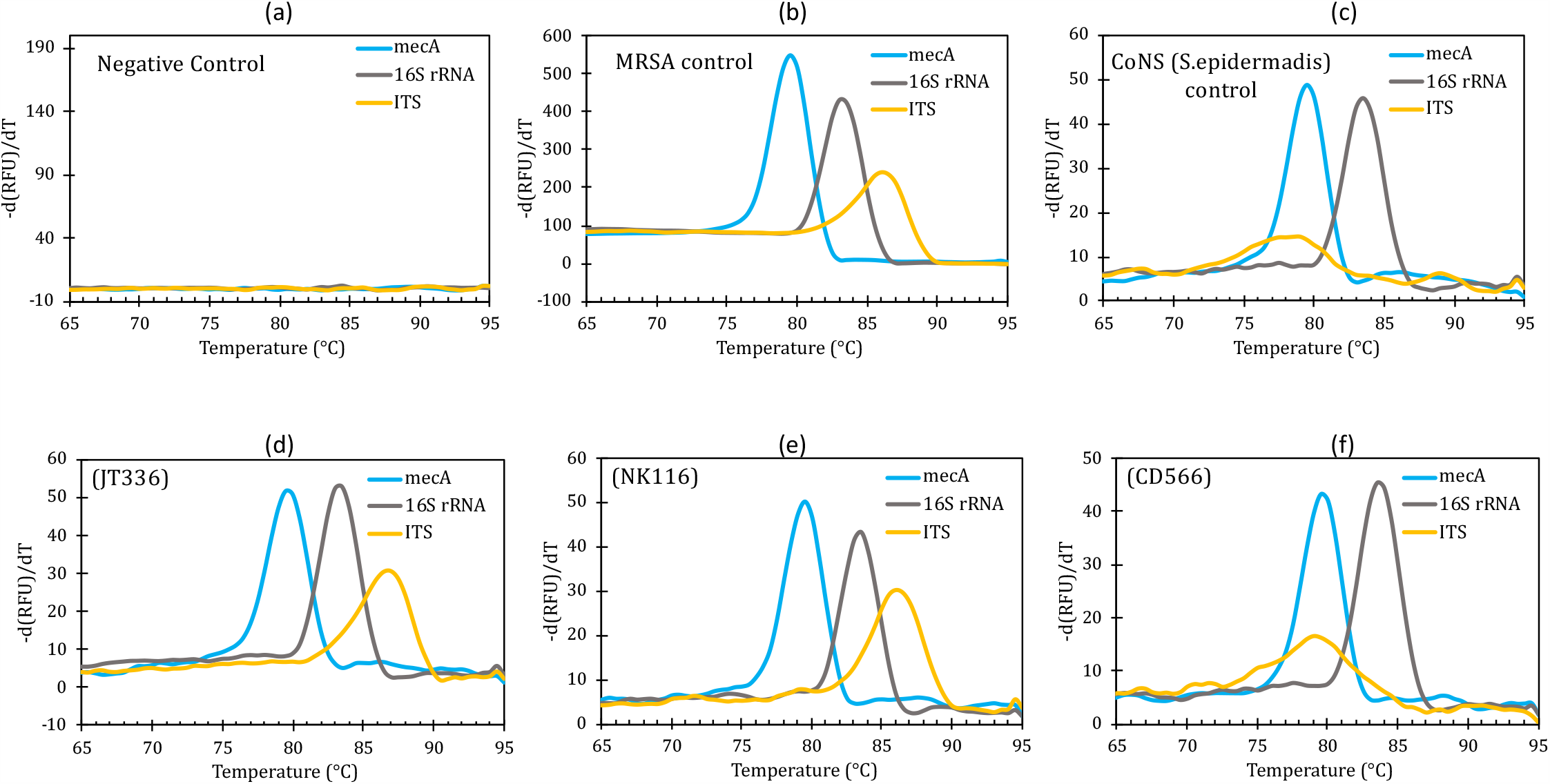
Melt curve analysis (MCA) for identifying CoNS contination in the discordant samples.(a) to (c) Melt curves for three control samples and (d) to (f) three representative samples (JT336, NK116 and CD566)

## 4. Discussion

Infection by MRSA is a widely acknowledged public health challenge as one of the hospital-acquired infections.(33) Low affinity penicillin-binding protein (PBP2), an altered protein encoded by the *mecA* gene present in the chromosome *Staphylococcal* cassette chromosome *mec* (SCCmec) is responsible for manifesting *methicillin* resistance in *S. aureus*.(34,35) Treatment of MRSA is often challenging due to its association with multiple antimicrobial resistances. Thus a rapid and accurate detection of MRSA infections can play a critical role in effective treatment and management of patients (36).

In conventional culture-based detection of MRSA, samples showing bacterial growth on primary plates are further processed for strain identification tests like coagulase and catalase and then finally for anti-microbial susceptibility using disc diffusion assay. Culture based detection of MRSA face inherent challenges including but not limited to longer processing time and limited range of detection in low inoculum culture (12). Thus, the need of a rapid and robust detection of MRSA has led to the development of multiple detection approaches, majorly based on nucleic acid amplification of a specific gene in MRSA genome (37,38). Despite the large number of efforts, a plethora of literature is available showcasing the shortcomings of the available detection methods and the resultant variation among the diagnostic methods (25,39).

MRSA detection can be performed on clinical isolates from several anatomical sites like wound exudates (pus), nasal swabs, skin swabs, and blood samples with nasal swabs being preferred by clinical test kits (40,41). However, the relevance of pus culture over other anatomic samples, especially blood, is recently reported in the literature with new tests being developed based on pus cultures (14,42,43).

The present study compared the diagnostic outputs of MRSA detection by three commonly used approaches namely, culture-based method, *q*PCR, and loop-mediated isothermal amplification. On comparing the diagnostic outputs of these methods, 95 samples were found to be negative across the three methods and thus taken as negative controls for the purpose of statistical analysis. A significant difference between the sensitivities of these methods was seen with the culture-based method detecting 30 MRSA positive samples while *q*PCR based assay detected 67 positive samples. Highest MRSA detection of 104 samples was observed in LAMP-based assay. It would be imperative to mention that MRSA presence in 39 samples were exclusively identified by LAMP method (Table 4a).

Both overall and pairwise discordance were estimated and suggested considerable differences in the diagnostics outputs(Figure 4a, 4b). Diagnostic results for NAATs methods (*q*PCR and LAMP) showed significant discordance with culture method, and were able to detect more number of MRSA bearing samples. The observed discordance between the diagnostic outputs of culture and molecular method highlights the limitation of traditional method given the time consuming protocols process and other variables including the slow rate of growth of bacteria, reduced sensitivity due to presence of other pathogens, and pus being a heterogeneous body fluid bearing high concentration of immune cells that hampered the viability of the MRSA pathogen (44). Results of NAAT methods including *q*PCR and LAMP were found to be more largely similar in comparison to culture based assay. Among the 69 samples found discordant between the NAATs methods, 53 were exclusively detected in LAMP while 16 were detected MRSA positive in qPCR method. Since both LAMP and qPCR assay are based on mecA gene amplification, observed discordance between the two is not well-understood. Although, it can be explained to some extent considering high sensitivity and robustness reported for LAMP based assays in multiple reports (32,45).

In addition to this, the presence of another pathogen CoNS possessing *mecA* gene and its influence on NAAT methods based detection cannot be overlooked (31). The methicillin resistance target gene *mecA* is often found in two organisms namely, CoNS and *S. aureus*,(28) and hence merely detecting *mecA* is not sufficient to distinguish between MRSA and methicillin resistant CoNS. Kahánková et al. employed a multi-locus PCR based approach to amplify multiple DNA segments of both species simultaneously (46). However, the performance of multi-locus PCR has yielded limited success as reported by other research groups (47,48). Hence, melt curve analysis was performed in the 69 clinical pus samples exhibiting discordant results to rule out the detection of false positives and differentiate between MRSA and CoNS (Table 5). Seventeen samples were found to be CoNS positive, while the rest 52 samples were found to be MRSA positive. Statistical analysis was again performed for NAATs methods while considering the CoNS identified samples as negative (Table 6). The results show that isothermal amplification (LAMP) based detection of MRSA was found to be the robust approach among the three methods, with sensitivity of 85.7 and NPV of 86.7% (Table 6).

**Table 6.** MRSA detection results and statistical analysis for LAMP and *q*PCR method after considering CoNS contamination.

| Nucleic acid based Diagnostic Methods | From melt curve analysis |  |  |  | Sensitivity (%) | Specificity (%) | Positive predictive value (PPV) (%) | Negative predictive value (NPV) (%) |
| --- | --- | --- | --- | --- | --- | --- | --- | --- |
|  | True Positive | True Negative | False positive* | False negative** |  |  |  |  |
| qPCR | 64 | 109 | 3 | 41 | 61.0 | 97.3 | 95.5 | 72.7 |
| LAMP assay | 90 | 98 | 14 | 15 | 85.7 | 87.5 | 86.5 | 86.7 |

*n*: number of samples; TP: true positive; FP: false positive; TN: true negative; FN: false negative; PPV: positive predictive value; NPV: negative predictive value; LAMP: loop-mediated isothermal amplification; qPCR: quantitative polymerase chain reaction.
\*False Positive – Samples detected as MRSA positive but found contaminated with CoNS through MCA.
\*\*False negative – (Total positive in the corresponding diagnostic method – True positive)

LAMP is comparatively the latest detection assay amongst the three methods and could be employed in hospital or commercial laboratory settings for rapid detection, given its low limit of detection and the capability to detect cell viability. Perhaps the best strategy for MRSA detection would be to use LAMP based assay for quick screening of clinical samples and then perform multiplex PCR or biochemical assay to re-confirm the presence of MRSA if so desired by the clinician. Such approach will help in saving the precious time lost with processing of negative samples in conventional methods, while reducing the cost too.

## 5. Conclusions

In conclusion, our study highlights significant discordance in outputs of the diagnostic performances and efficiency between three different methods used for MRSA detection in clinical pus isolates. Significant discordance was seen among the diagnostic results of three methods, with LAMP based method detecting highest number of MRSA infections. Within the nucleic acid amplification-based methods, case-wise comparison was made to identify the non-concordant samples. A melt curve analysis (MCA) was used to identify the presence of CoNS for non-concordant samples while other reasons leading to discrepancies are elaborately discussed. The study highlights the robustness and loopholes of different approaches used for MRSA method and the factors which should be considered for the development of an advanced and more specific detection method. One of the best strategies for MRSA detection would be to use LAMP based assay for quick screening of clinical samples and then perform multiplex PCR or biochemical assay to re-confirm the MRSA presence if desired by the clinician.

## Data Availability

All the data pertaining to the manuscript is available with authors.

## References

1. Kluytmans J, Harbarth S. MRSA transmission in the community: emerging from under the radar. Lancet Infect Dis. 2019;

2. Dulon M, Haamann F, Peters C, Schablon A, Nienhaus A. Mrsa prevalence in european healthcare settings: A review. BMC Infect Dis. 2011;

3. Otto M. Community-associated MRSA: What makes them special? International Journal of Medical Microbiology. 2013.

4. Pierce R, Lessler J, Popoola VO, Milstone AM. Meticillin-resistant Staphylococcus aureus (MRSA) acquisition risk in an endemic neonatal intensive care unit with an active surveillance culture and decolonization programme. J Hosp Infect. 2017;95(1):91–7.

5. Coughenour C, Stevens V, Stetzenbach LD. An evaluation of methicillin-resistant Staphylococcus aureus survival on five environmental surfaces. Microb drug Resist. 2011;17(3):457–61.

6. van de Sande-Bruinsma N, Leverstein van Hall MA, Janssen M, Nagtzaam N, Leenders S, de Greeff SC, et al. Impact of livestock-associated MRSA in a hospital setting. Antimicrob Resist Infect Control. 2015;

7. Zetola N, Francis JS, Nuermberger EL, Bishai WR. Community-acquired meticillin-resistant Staphylococcus aureus: An emerging threat. Lancet Infectious Diseases. 2005.

8. Goh ZNL, Chung PY. Letters to the Editor Incidence of meticillin-resistant Staphylococcus aureus contamination on mobile phones of medical students Is it necessary to test the sterility of urine prior to outpatient cystoscopy ? J Hosp Infect. 2019;101(4):482–3.

9. Bellini D, Koekemoer L, Newman H, Dowson CG. Novel and Improved Crystal Structures of H. influenzae, E. coli and P. aeruginosa Penicillin-Binding Protein 3 (PBP3) and N. gonorrhoeae PBP2: Toward a Better Understanding of β-Lactam Target-Mediated Resistance. J Mol Biol. 2019;

10. Østergaard C, Møller JK. Subdivision of MRSA CC398 isolates using MALDI-TOF MS. Int J Med Microbiol. 2018;

11. Cadena J, Thinwa J, Walter EA, Frei CR. Risk factors for the development of active methicillin-resistant Staphylococcus aureus (MRSA) infection in patients colonized with MRSA at hospital admission. Am J Infect Control. 2016;44(12):1617–21.

12. Kniehl E, Becker A, Forster DH. Bed, bath and beyond: Pitfalls in prompt eradication of methicillin-resistant Staphylococcus aureus carrier status in healthcare workers. J Hosp Infect. 2005;

13. Silverman SM, Moses JE, Sharpless KB. Reengineering Antibiotics to Combat Bacterial Resistance: Click Chemistry [1,2,3]-Triazole Vancomycin Dimers with Potent Activity against MRSA and VRE. Chem - A Eur J. 2017;

14. Kaur A, Ruhela A, Sharma P, Khariwal H, Seth S, Kumar A, et al. Simultaneous and high sensitive detection of Salmonella typhi and Salmonella paratyphi a in human clinical blood samples using an affordable and portable device. Biomed Microdevices. 2019;

15. Abram TJ, Cherukury H, Ou C-Y, Vu T, Toledano M, Li Y, et al. Rapid bacterial detection and antibiotic susceptibility testing in whole blood using one-step, high throughput blood digital PCR. Lab Chip. 2020;

16. Paule SM, Pasquariello AC, Thomson RB, Kaul KL, Peterson LR. Real-time PCR can rapidly detect methicillin-susceptible and methicillin-resistant Staphylococcus aureus directly from positive blood culture bottles. Am J Clin Pathol. 2005;

17. McElhinney R, Millar C, Scopes E. Comparative evaluation of chromID MRSA agar and Brilliance 2 MRSA agar for detection of MRSA in clinical samples. Br J Biomed Sci. 2013;

18. Eigner U, Veldenzer A, Holfelder M. Validation of the FluoroType® MRSA assay for the rapid identification of methicillin-resistant Staphylococcus aureus directly from patient material. J Microbiol Methods. 2014;

19. Notomi T, Okayama H, Masubuchi H, Yonekawa T, Watanabe K, Amino N, et al. Loop-mediated isothermal amplification of DNA. Nucleic Acids Res. 2000;28(12):E63.

20. Medalla F, Gu W, Mahon BE, Judd M, Folster J, Griffin PM, et al. Estimated incidence of antimicrobial drug--resistant nontyphoidal Salmonella infections, United States, 2004--2012. Emerg Infect Dis. 2017;23(1):29.

21. Eriksson E, Aspan A. Comparison of culture, ELISA and PCR techniques for salmonella detection in faecal samples for cattle, pig and poultry. BMC Vet Res. 2007;3.

22. Tanner NA, Zhang Y, Evans TC. Visual detection of isothermal nucleic acid amplification using pH-sensitive dyes. Biotechniques. 2015;

23. Tomita N, Mori Y, Kanda H, Notomi T. Loop-mediated isothermal amplification (LAMP) of gene sequences and simple visual detection of products. Nat Protoc. 2008;

24. Tansarli GS, LeBlanc L, Auld DB, Chapin KC. Diagnostic accuracy of pre-surgical Staphylococcus aureus PCR assay compared to culture and post-PCR implementation surgical site infection rates. J Mol Diagnostics [Internet]. 2020 May [cited 2020 May 31];0(0). Available from: https://linkinghub.elsevier.com/retrieve/pii/S1525157820303275

25. Rabaan AA, Bazzi AM. Variation in MRSA identification results from different generations of Xpert MRSA real-time PCR testing kits from nasal swabs. J Infect Public Health. 2017;

26. Alipour F, Ahmadi M, Javadi S. Evaluation of different methods to detect methicillin resistance in Staphylococcus aureus (MRSA). J Infect Public Health. 2014;

27. Gunderson CG, Holleck JL, Chang JJ, Merchant N, Lin S, Gupta S. Diagnostic accuracy of methicillin-resistant Staphylococcus aureus nasal colonization to predict methicillin-resistant S aureus soft tissue infections. Am J Infect Control. 2016;44(10):1176–7.

28. Bowers KM, Wren MWD, Shetty NP. Screening for methicillin resistance in Staphylococcus aureus and coagulase-negative staphylococci: An evaluation of three selective media and Mastalex-MRSA latex agglutination. Br J Biomed Sci. 2003;

29. CLSI document M47-A. Principles and Procedures for Blood Cultures; Aproved Guideline. Clin Lab Standars Inst. 2007;

30. Notomi T, Mori Y, Tomita N, Kanda H. Loop-mediated isothermal amplification (LAMP): principle, features, and future prospects. 2015;53(1):1–5.

31. Heydari N, Alikhani MY, Tahmasebi H, Asghari B, Arabestani MR. Design of melting curve analysis (MCA) by real-time polymerase chain reaction assay for rapid distinction of staphylococci and antibiotic resistance. Arch Clin Infect Dis. 2019;

32. Lee D, Kim EJ, Kilgore PE, Kim SA, Takahashi H, Ohnishi M, et al. Clinical Evaluation of a Loop-Mediated Isothermal Amplification (LAMP) Assay for Rapid Detection of Neisseria meningitidis in Cerebrospinal Fluid. PLoS One. 2015;10(4):e0122922.

33. Deurenberg RH, Vink C, Kalenic S, Friedrich AW, Bruggeman CA, Stobberingh EE. The molecular evolution of methicillin-resistant Staphylococcus aureus. Clinical Microbiology and Infection. 2007.

34. Brukner I, Oughton M, Giannakakis A, Kerzner R, Dascal A. Significantly improved performance of a multitarget assay over a commercial sccmec-based assay for methicillin-resistant staphylococcus aureus screening: Applicability for clinical laboratories. J Mol Diagnostics. 2013 Sep 1;15(5):577–80.

35. Ito T, Katayama Y, Asada K, Mori N, Tsutsumimoto K, Tiensasitorn C, et al. Structural comparison of three types of staphylococcal cassette chromosome mec integrated in the chromosome in methicillin-resistant Staphylococcus aureus. Antimicrob Agents Chemother. 2001;

36. Grundmann H, Aires-de-Sousa M, Boyce J, Tiemersma E. Emergence and resurgence of meticillin-resistant Staphylococcus aureus as a public-health threat. Lancet. 2006.

37. Becker K, Denis O, Roisin S, Mellmann A, Idelevich EA, Knaack D, et al. Detection of mecA-and mecC-positive methicillin-resistant staphylococcus aureus (MRSA) isolates by the new Xpert MRSA Gen 3 PCR assay. J Clin Microbiol. 2016;

38. Palavecino EL. Rapid methods for detection of MRSA in clinical specimens. Methods Mol Biol. 2014;

39. Chan C, Carson L, Smith GC, Morelli A, Lee S. Applied Surface Science Enhancing the antibacterial performance of orthopaedic implant materials by fibre laser surface engineering. Appl Surf Sci. 2017;404:67–81.

40. Peterson LR, Woods CW, Davis TE, Wang ZX, Young SA, Osiecki JC, et al. Performance of the cobas MRSA/SA Test for Simultaneous Detection of Methicillin-Susceptible and Methicillin-Resistant Staphylococcus aureus from Nasal Swabs. Am J Clin Pathol. 2017;

41. Grmek-Kosnik I, Dermota U, Ribic H, Storman A, Petrovic Z, Zohar-Cretnik T. Evaluation of single vs pooled swab cultures for detecting MRSA colonization. J Hosp Infect. 2018;98(2):149–54.

42. Sloane AJ, Pressel DM. Culture Pus, Not Blood: Decreasing Routine Laboratory Testing in Patients With Uncomplicated Skin and Soft Tissue Infections. Hosp Pediatr. 2016;

43. Zwemer E, Stephens JR. Things we do for no reason: Blood cultures for uncomplicated skin and soft tissue infections in children. J Hosp Med. 2018;

44. Rudkjøbing VB, Thomsen TR, Xu Y, Melton-Kreft R, Ahmed A, Eickhardt S, et al. Comparing culture and molecular methods for the identification of microorganisms involved in necrotizing soft tissue infections. BMC Infect Dis. 2016;

45. Yang H, Ma X, Zhang X, Wang Y, Zhang W. Development and evaluation of a loop-mediated isothermal amplification assay for the rapid detection of Staphylococcus aureus in food. Eur Food Res Technol. 2011;

46. Kahánková J, Pantůček R, Goerke C, Růžičková V, Holochová P, Doškař J. Multilocus PCR typing strategy for differentiation of Staphylococcus aureus siphoviruses reflecting their modular genome structure. Environ Microbiol. 2010;

47. Stürenburg E. Rapid detection of methicillin-resistant Staphylococcus aureus directly from clinical samples: methods, effectiveness and cost considerations. Germanmedical science : GMS e-journal. 2009.

48. French GL. Methods for screening for methicillin-resistant Staphylococcus aureus carriage. Clinical Microbiology and Infection. 2009.

